# Validation of an innovative two-part algorithm for detecting self-propulsion in manual wheelchair users

**DOI:** 10.1101/2024.11.14.24313548

**Authors:** Rose Gagnon, Krista L. Best, François Routhier

## Abstract

**Introduction:** Actimetry is increasingly used to measure physical activity (PA) for MWC users. However, conversion of raw data into interpretable PA outcomes remains imprecise, and the differentiation between propulsion and non-propulsion is challenging. Using a previously developed two-part algorithm, the objectives of this study were to 1) measure the accuracy of total distance collected, and 2) validate the algorithm’s accuracy in differentiating between self-propulsion and non-propulsion.

**Methods:** Experimental study consisting of two data collection sessions. Actimetry data (Actigraph) were collected indoors (controlled conditions) during 100 repetitions (n=40 MWC propulsion, n=60 pushing the MWC) over three distances (10, 50 and 100 meters). Actimetry data (Actigraph) were also collected outdoors (uncontrolled condition) during self-propulsion over 1,000 meters (10 repetitions). Descriptive statistics (e.g., mean, standard deviation) with confidence intervals and accuracy measures (percentage of true value) were conducted for each trial.

**Results:** The algorithm measured total distance covered indoors with an excellent accuracy (98.9% to 99.8%). It differentiated between self-propulsion and non-propulsion with an accuracy between 96.2% and 99.2% under controlled condition, and between 91.3% and 100.0% under uncontrolled condition.

**Conclusions:** The algorithm tested allowed precise measurement of total distance covered, as well as an excellent discrimination between self-propulsion and non-propulsion.

## Introduction

Physical activity (PA) guidelines recommend at least 150 minutes of moderate to vigorous PA per week to maintain good health.^1^ Although this recommendation applies to people who use manual wheelchairs (MWC), barriers such as time and energy, physical health, accessibility of sports facilities, and transportation restrict attainment of this goal.^2, 3^ Furthermore, the variability of health conditions and disabilities among people who use MWC makes the development of a single PA measure very complex.^4^ While more and more technologies are being developed to measure PA levels in ambulatory populations (e.g., cellphone applications, smartwatches), their adaptation to the reality of MWC use is still in its early stages.^5^ However, recent studies (e.g., ^6, 7^) report that the use of accelerometers could be a promising avenue for measuring PA in MWC users.

Accelerometers are discrete devices that accurately characterize movement,^8, 9^ making them a method of choice for objectively and validly measuring PA levels and intensity. Since they are small and lightweight, wearing them does not impede body movements.^10^ Accelerometers offer many benefits in ambulatory populations, including monitoring of PA and related goals, increased motivation to perform PA, and built-in feedback.^11-13^ They also enable PA to be measured more easily and effectively in uncontrolled environments.^14^ For all these reasons, recent years have seen the development and adaptation of several types of accelerometers for measuring PA in MWC users, such as SenseWear,^15^ ReSense,^16^ Apple Watch,^17^ activPAL,^18^ PAMS^19, 20^ or Actiwatch^21^. The most widely used accelerometer, however, remains the actigraph, such as the Actigraph GT3X+, given its validated accuracy, reliability, comfort and ability to measure energy expenditure and different types of PA.^6, 10, 22-24^ Actigraphs can be attached to the wheel or worn on the arm^25^ to collect tri-axial data during the wheel revolutions, absolute angle and duration of movement, which can be transformed into objective measures of mobility (i.e., total distance travelled, number of bouts, speed and duration of movement).^4, 6, 8^ Actigraphs can also provide estimates of PA frequency, intensity and duration of an activity.^5^ However, the ability to distinguish between MWC propulsion and non-propulsion (i.e., being pushed by someone else) with actigraphs remains a challenge to this day.^7, 26^ Furthermore, to date, actigraph validation has oftentimes been conducted in relatively controlled environments that are not representative of everyday life (e.g., laboratory setting, hard wooden surface such as a gymnasium).^9, 27, 28^ Moreover, the algorithms used to convert raw actimetry data into PA measurements are: 1) often unable to derive a distance traveled or a speed,^10^ 2) have poor validity for measuring small-amplitude movements or speeds,^27^ 3) are not very or not transparent at all,^10, 27-30^ or 4) are unable to differentiate between self-propulsion and passive pushing of the MWC^8, 10, 18, 21, 27^.

In an attempt to address these challenges, our team has developed a two-part custom algorithm for calculating total distance travelled in a MWC, as well as discriminating between propulsion and non-propulsion. Using this two-part algorithm, the objectives of this study were to 1) measure the accuracy of total distance collected in a controlled setting (indoors), and 2) validate the algorithm’s accuracy in differentiating between self-propulsion and non-propulsion in controlled (indoors) and uncontrolled settings (outdoors).

## Methods

### Study design

This study was an experimental study aimed at validating a two-part algorithm that will be used to quantify MWC users’ activity level and overall mobility.^31^ Data collection was completed during two, 1.5 hour sessions at a university campus and affiliated research centre. The *Centre Intégré Universitaire de Santé et de Services Sociaux de la Capitale-Nationale* Research Ethics Board in rehabilitation and social integration waived the ethical approval requirement for this study (#2025-32390).

### Data collection

An Actigraph wGT3X-BT was used for all data collections (see Table 1 for a summary of characteristics).

**Table 1.** Actigraph wGT3X-BT characteristics^32^.

|  |  |
| --- | --- |
| **Weight** | 19 grams |
| **Dimensions** | Length: 4.6 cm, Width: 3.3 cm, Height: 1.5 cm |
| **Sample Rate** | 30 to 100 Hertz |
| **Data Storage** | 180 days |
| **Battery Life** | Greatly varies, but between 3 to 5 weeks for this application |
| **Data Transfer** | USB and Bluetooth |
| **Where to Wear** | Wrist, waist, ankle and thigh |

The Actigraph was worn on the upper arm (and not on the wrist) to prevent interference with the natural movement of MWC propulsion and since the measurement accuracy is almost identical between the two locations.^33^ Data was transferred onto a computer using the Actilife software (ActiGraph, 2023)^34^ and was processed using the two-part algorithm. The MWC used during the two data collections was a Quickie GPV.^35^

### Indoor controlled setting

The first data collection took place in the underground tunnels of a university campus by a study investigator. Actigraph data was collected over short (10 meters) and medium (50 and 100 meters) distances in two phases (propulsion, pushing the MWC). First, while sitting in the MWC, the study investigator propelled the MWC over each distance using two arms (n=40 repetitions). Actigraphs were worn on the upper arm and on one rear wheel of the MWC. Second, the study investigator pushed the MWC over each distance (n=60 repetitions), and an actigraph was placed only on the rear wheel of the MWC.

The Gold Standard used to measure the linear distance traveled during this first data collection was a DBV50 Core measuring wheel encoder.^36^ The device can be placed at the back of a wheelchair and is connected to a tablet or computer that displays the distance. Table 2 summarizes its main characteristics.

**Table 2.** DBV50 Core measuring wheel encoder characteristics^37^.

|  |  |
| --- | --- |
| **Pulse per revolution** | 2 500 (10 000 pulse per turn in quadrature) |
| **Resolution** | 12.5 pulses per mm and 0.08 mm per pulse |
| **Measuring step deviation** | $\pm 18^\circ$ per pulses per revolution |
| **Error limits** | $\pm 4$ mm/m (influenced by the measuring wheel) |
| **Duty cycle** | $\leq 0.5 \pm 5\%$ |
| **Initialization time** | Less than 3 ms |

The wheel encoder was used as a standard measurement over all distances (10, 50 and 100m) to calibrate the algorithm. The USB interface used was a PhidgetEncoder HighSpeed.^38^ This interface was used to keep track of MWC movement.

### Outdoor uncontrolled setting

Ten members of the research team participated in the second data collection (1 1,000-meter data collection per team member). To be part of the validation process, team members had to have had some experience in the use of a MWC before and to be able to propel themselves over 1,000 meters. Data collection took place on a winding bicycle path with a few slopes and varying terrain (e.g., grass, pavement) near the university affiliated research centre. To measure arm and MWC movement in a real-life context with the actigraphs, team members self-propelled a MWC at a self-selected pace over 1,000 meters. Two actigraphs were worn, one on the upper arm and one on the rear wheel of the MWC. The high-resolution encoder could not be used as a standard measurement for the second data collection due to the irregular and rough surface of the path (i.e., the vibration created by the surface caused the measured distance to vary too much). Therefore, a measuring wheel similar to the Lufkin 12” Professional Metric Measuring Wheel^39^ was used.

### Data processing

Raw data were processed using a custom algorithm (MATLAB 2019b, MathWorks)^40^ with two distinct parts: 1) analyze the distance traveled by the rear wheel of the MWC (wheelchair activity algorithm) and 2) differentiate between self-propulsion in the MWC and the MWC being pushed (autonomous propulsion detection algorithm) (Figure 1).

**Figure 1.**
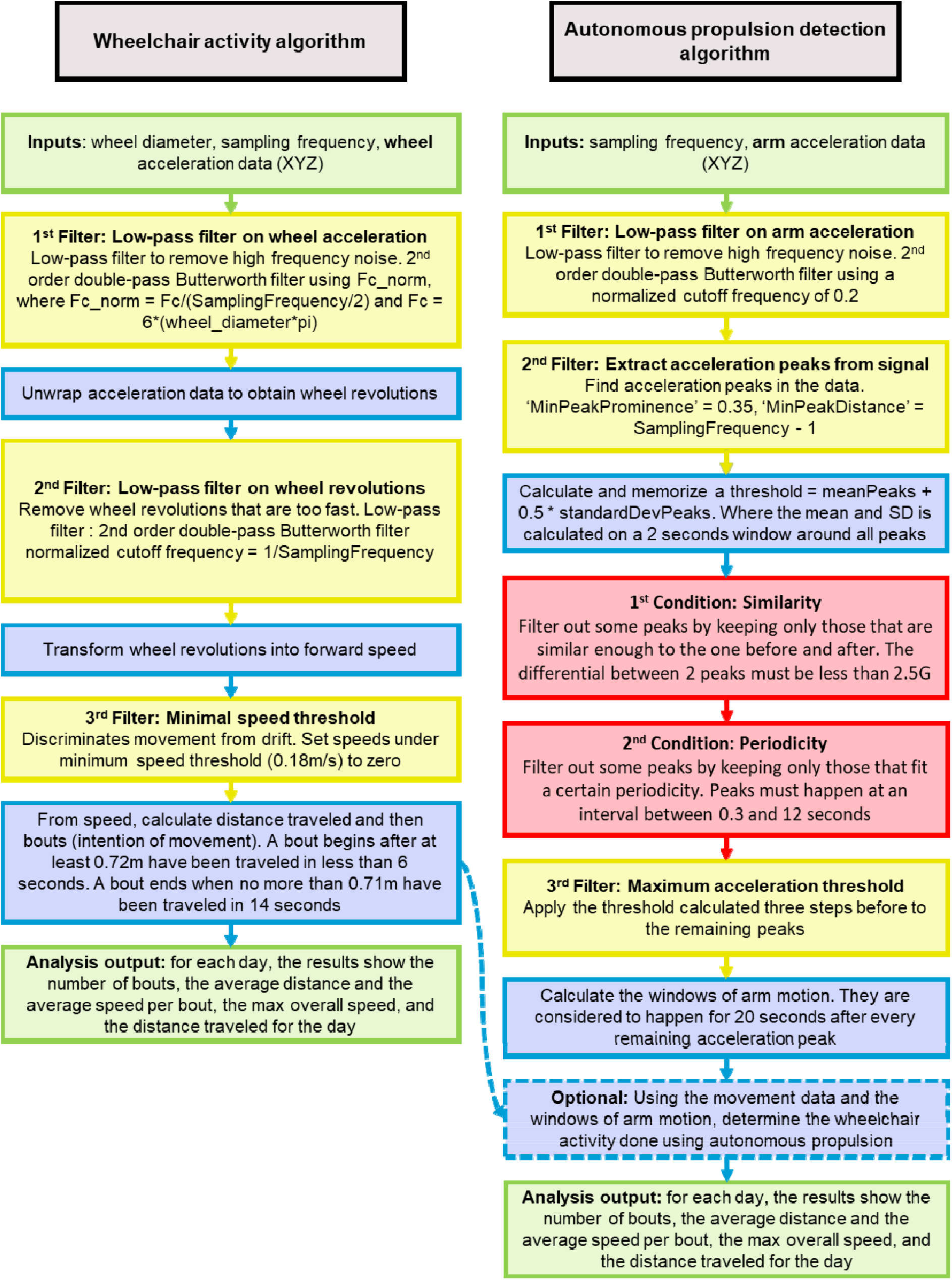
Detail of the two-part algorithm for calculating the distance traveled by the MWC and the presence of propulsion

The two parts of the algorithm required the following information to analyze data correctly: 1) the wheelchair wheel diameter, 2) the sampling frequency, and 3) the three acceleration vectors from the actigraph (X, Y and Z) (Figure 1). To be considered as motion, the signal analyzed in the two parts of the algorithm passed through a series of filters acting on the frequency domain. Acceleration peaks that were too far apart or too close together were removed to retain only those representing continuous motion that can be achieved by a person propelling themself in a MWC.

A series of two conditions were applied to the acceleration peaks in the self-propulsion detection algorithm to ensure that the amplitude was within the expected values (Figure 1). If a peak failed to meet the first condition, it was rejected and did not pass to the second, until all peaks were passed through the two conditions (Figure 1). Once the acceleration data was transformed into speed within the wheelchair activity algorithm, an optional function was called to remove from the distance vector the movement that occurred when the arm was motionless. If nothing was specified, the algorithm calculated the total distance traveled by the rear wheel regardless of how the distance was traveled. If necessary, the algorithm divided the analysis output in 24-hour windows as the code was written with long data collection sessions in mind (i.e., to facilitate data collection in real-world settings).

The MATLAB programming required for the wheelchair activity algorithm and the filters were based on the work of Sonenblum et al.^41^ The minimal wheel movement to consider a movement as a bout (0.72 m) came from the work of Tolerico et al. who demonstrated that the average minimal speed of movement of MWC users with various health conditions in the home was 0.72 m/s.^4^ All the parameters and filters of the autonomous propulsion detection algorithm were developed by our research team based on the characteristics of arm movements listed in the literature for a population of walkers in whom data were taken with an actigraph. Finally, the two conditions used were based on the work of Gu et al.^42^

### Data analysis

All accelerations from the actigraph data collections (10, 50, 100, and 1,000 meters) were processed using the two-part algorithm presented earlier and are presented as distances traveled in meters. Since our sample size was small (n=100 for all distances except the 1,000 meters where n=10), we bootstrapped (n=1,000) data obtained from the algorithm for each of the measured distances to obtain robust descriptive statistics (mean, standard deviation, 95% confidence interval) for each distance assessed.^43^ Accuracy measures (percentage of true value) were also conducted for each trial. All statistical analyses were performed using Excel (Microsoft 365, 2011) and SPSS statistical software (Version 25; IBM, Armonk, NY, USA).

## Results

The distances measured at the wheel and arm when the MWC was propelled in a controlled environment are shown in Table 3:

**Table 3.** Measurements of the distance covered by the wheel of the wheelchair and the arm when propelling obtained using the algorithm.

| Trial type | Number of included trials | Mean | Standard deviation | Accuracy | Sample's 95% confidence interval |  | 95% bootstrapped confidence interval |  |
| --- | --- | --- | --- | --- | --- | --- | --- | --- |
|  |  | m | m | % | m |  | m |  |
| **10 meters – Wheel** | 30 | 9.7363 | 0.45605 | 97.36 | 9.5660 | 9.9066 | 9.5787 | 9.8833 |
| **10 meters – Arm** | 30 | 9.6193 | 0.51276 | 96.19 | 9.4279 | 9.8108 | 9.4237 | 9.7980 |
| **50 meters – Wheel** | 40 | 50.3809 | 0.43354 | 99.24 | 50.2422 | 50.5195 | 50.2377 | 50.5113 |
| **50 meters – Arm** | 40 | 49.5758 | 1.09675 | 99.15 | 49.2250 | 49.9265 | 49.2063 | 49.8859 |
| **100 meters – Wheel** | 40 | 101.6318 | 1.16632 | 98.37 | 101.2587 | 102.0048 | 101.2998 | 101.9912 |
| **100 meters – Arm** | 40 | 98.6595 | 7.74597 | 98.66 | 96.1822 | 101.1368 | 96.0182 | 100.7193 |

m: meters, %: percentage

The distance measurements obtained at the rear wheel are lower than the reference distance for the 10-meter propulsion trials, whereas they are higher for the 50-meter and 100-meter propulsion trials. Furthermore, although the observed and calculated confidence intervals oscillate close to the reference values, they do not contain any of the expected distance values (10, 50 and 100 meters). The same applies to distance measurements derived via the arm’s actigraph. The observed and calculated confidence intervals of the 10- and 50-meter tests are close to, but do not overlap, the reference distance. The two confidence intervals for the 100-meter tests nevertheless include the reference value. Despite the variability observed in the confidence intervals, the observed accuracy remains over 96%, for both arm and rear wheel measurements.

Distances measured at the rear wheel when the MWC was pushed in a controlled environment and calculated using the two-part algorithm are presented in Table 4:

**Table 4.**
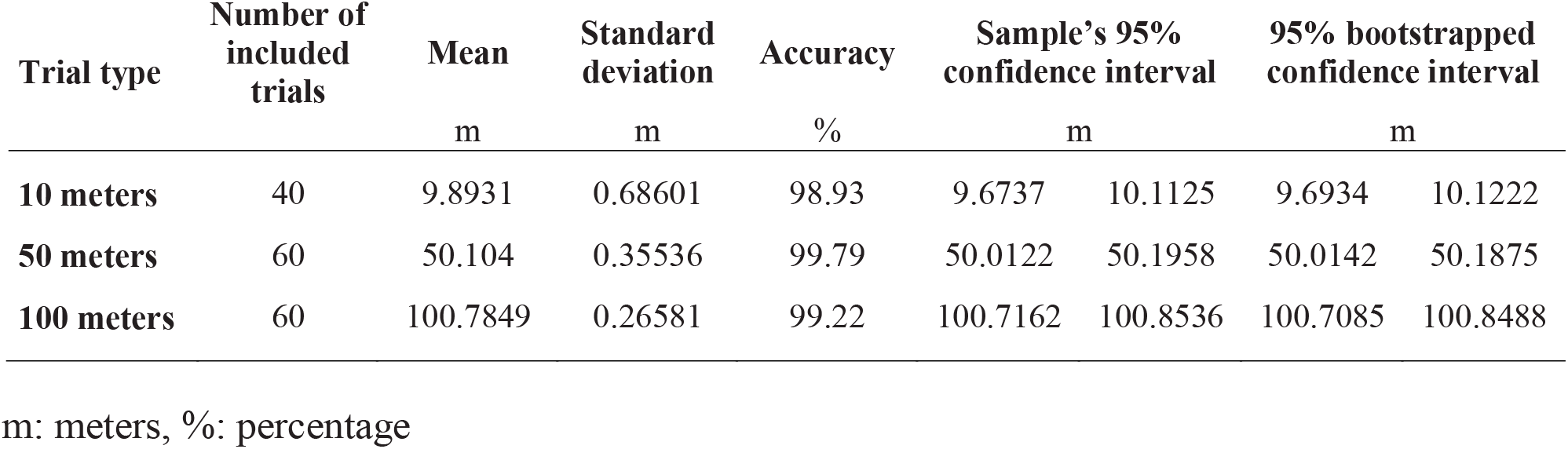
Measurements of the distance covered by the wheel of the wheelchair when pushed obtained using the algorithm.

| Trial type | Number of included trials | Mean | Standard deviation | Accuracy | Sample's 95% confidence interval |  | 95% bootstrapped confidence interval |  |
| --- | --- | --- | --- | --- | --- | --- | --- | --- |
|  |  | m | m | % | m |  | m |  |
| **10 meters** | 40 | 9.8931 | 0.68601 | 98.93 | 9.6737 | 10.1125 | 9.6934 | 10.1222 |
| **50 meters** | 60 | 50.104 | 0.35536 | 99.79 | 50.0122 | 50.1958 | 50.0142 | 50.1875 |
| **100 meters** | 60 | 100.7849 | 0.26581 | 99.22 | 100.7162 | 100.8536 | 100.7085 | 100.8488 |

m: meters, %: percentage

Although the mean of the 10-meter trials is slightly less than 10 meters, the 95% confidence interval of the sample and that obtained from the Bootstrap analyses include the target measurement. Conversely, the mean distances calculated by the algorithm for the 50- and 100-meter trials are higher than the reference distances and are not included in either the samples’ confidence intervals or those obtained by bootstrapping. Nevertheless, the accuracy of the algorithm remains close to 100% for all trial types.

Table 5 shows data from the 10 team members who propelled themselves in the MWC over 1,000 meters in a real-life setting:

**Table 5.** Distance travelled and measured by the algorithm during propulsion over one kilometer in uncontrolled conditions.

| **Participant** | **Travelled distance**<br>m | **Measured distance**<br>m | **Accuracy**<br>% |
| --- | --- | --- | --- |
| **False positive 1** | 709.0 | 0.0 | 100.00 |
| **False positive 2** | 615.0 | 40.0 | 93.49 |
| **False positive 3** | 669.0 | 0.0 | 100.00 |
| **Team member 1** | 1022.2 | 1022.2 | 100.00 |
| **Team member 2** | 1021.7 | 1020.6 | 99.89 |
| **Team member 3** | 1017.0 | 1007.3 | 99.05 |
| **Team member 4** | 1026.0 | 1013.2 | 98.75 |
| **Team member 5** | 1027.0 | 1006.8 | 98.03 |
| **Team member 6** | 1023.0 | 934.3 | 91.33 |
| **Team member 7** | 1017.0 | 979.9 | 96.35 |
| **Team member 8** | 1022.9 | 1017.4 | 99.46 |
| **Team member 9** | 1023.0 | 1020.8 | 99.78 |
| **Team member 10** | 1016.0 | 988.8 | 97.33 |
| **Mean accuracy** |  |  | **97.96** |

m: meters, %: percentage

The distance data calculated with the algorithm are very close to the expected values, except in the case of False Positive 2 and Team member 6, where a larger difference was observed. Despite this variability, the accuracy of the algorithm remains over 91%, with several distance measurements approaching an accuracy of 100% (mean: 97.96%).

## Discussion

Using a publicly available software (MATLAB), we previously developed a two-part algorithm for measuring total distance covered and differentiating between MWC propulsion and non-propulsion. The algorithm was designed to be used to derive a range of movement data, such as total distance travelled, average and maximum speeds, and number of bouts made, while offering the possibility of separating the output into 24-hour windows to facilitate the interpretation of data collected over long periods. The results of our study demonstrated that the developed two-part algorithm offers excellent accuracy, ranging from 96.2% to 99.8% for indoor movement measurements, and close to 98% in uncontrolled condition (outdoors).

Several other authors, such as Popp et al.^16^ and Callupe Luna et al.^44^ have also worked on algorithms to discriminate between propulsion and non-propulsion in the context of MWC use. Although they achieved an accuracy similar to our own (93% and 94%, respectively), this was achieved using four sensors (Popp et al.) and an invasive measurement system (Callupe Luna et al.), which had the potential to impede and even modify the natural propulsive motion.^45^ The trials used to validate their algorithms were also carried out in a controlled environment. Our study addresses these limitations, in that our algorithm enables precise measurement using just two sensors. We also carried out trials in an uncontrolled environment to test the algorithm in real-life conditions. Recent years have also seen the emergence of algorithms based on deep machine learning to categorize movements performed while using a MWC. Unlike other algorithms, these automatically generate the most optimal parameters for processing actimetry data.^46^ Although interesting levels of accuracy have been obtained in these studies (Van Der Slikke et al.: 83-89%,^47^ Fortune et al.: 88-100%,^48^ Garcia-Masso et al.: 61-94%,^49^ Hiremath et al.: 93-94%,^15^ De Vries et al.: >98%^46^), much remains to be done to democratize the use of this technology. Indeed, all these studies were carried out under controlled conditions (e.g., laboratory), with participants who had been trained to perform the MWC’s tasks. In addition, one of these studies used a large number of accelerometers,^49^ which could hinder natural propulsion movement, and another used an off-the-shelf activity measurement monitor.^15^ The integration of deep machine learning into our two-part algorithm could be of interest to optimize the selection of parameters to be considered when sorting actimetry data. Nevertheless, we believe that our algorithm in its present form makes an important contribution to the current actimetry literature, in that it uses data from a readily available accelerometer (Actigraph wGT3X-BT) and has excellent accuracy even under uncontrolled conditions.

The validation tests carried out in this study were performed by able-bodied team members with no particular disability. While these preliminary tests are important to ensure the algorithm’s reliability, subsequent validation tests should include both inexperienced and experienced MWC users, as they do not propel themselves in the same way.^26^ Similarly, it will be important to test the algorithm on populations with a diversity of health conditions to ensure its external validity.^7^

Moreover, the present two-part algorithm has been tested with actigraphs worn on the wheel of the wheelchair and on the upper limb. This configuration may not be suitable for all existing propulsion patterns (e.g., one foot and one arm, two feet). Further studies will be needed to adapt the data collection method to the reality of users propelling themselves without their upper limbs.^26, 47, 50^ As MWC users are at greater risk of developing various comorbidities given their generally greater physical inactivity,^51^ data collection using actigraphs has the potential to provide important objective clinical data when planning interventions to increase their PA practice.^26^ Greater use of actimetry data could also enable MWC users to monitor their health goals more closely, in addition to democratizing access to their PA data.^26^ Actimetry could also offer MWC users living in remote locations the possibility of having easier access to evidence-based health interventions.^51^

This study has a number of limitations. Firstly, the data for the 10-, 50- and 100-meter trials were collected in succession. Thus, all the 10-meter trials were carried out subsequently; the same method was used for all the 50- and 100-meter trials. This data collection method may have had two effects on collected data. Firstly, it may predispose to the presence of a learning effect throughout data collection. This method is also less representative of everyday movements, where wheelchair bouts’ duration and length are not necessarily predicted. In a second validation phase, it will be important to alternate the trials’ distance, as well as to validate the algorithm’s accuracy with data from trials where movements are initiated spontaneously. Furthermore, the same MWC was used for all trials. Subsequent studies should ensure that the algorithm’s reliability does not vary regardless of the MWC’s dimensions. Despite these limitations, this study has several strengths that are worth mentioning. The parameters used to create the conversion algorithm, and the functioning of the algorithm itself, are transparently reported. Distance data collected during all the trials were compared with distance measurements obtained using recognized standard measurements. The accuracy of the two-part algorithm was also evaluated both under controlled laboratory conditions and in an uncontrolled environment more representative of everyday activities. Finally, the algorithm presented can be used to calculate the total distance travelled in a MWC, as well as the total distance travelled using propulsion alone, in addition to deriving several other data characterizing wheelchair movements (i.e., number of bouts, average bout length, average and maximum speed).

## Conclusion

The present work represents a further step towards a better understanding and characterization of the PA performed by MWC users. Our algorithm was able to derive measures of total distance travelled, as well as distance travelled solely by propulsion. It proved accurate over a variety of distances (i.e., 10, 50, 100 and 1,000 meters), both in controlled and uncontrolled conditions. Further studies will be needed, however, to validate the algorithm in the context of spontaneous movements, which are more representative of everyday life. It will also be important to ensure that the algorithm remains valid for MWC users presenting a variety of health conditions and propulsion patterns. Better-quality PA measures have the potential to enhance the tailoring of clinical interventions recommended to MWC users.

## Data Availability

All data produced in the present study are available upon reasonable request to the authors.

## Notes

**Conflicting interests:** The Authors declare that there is no conflict of interest.

**Funding:** No external funding was received for this study. Krista Best and Francois Routhier received salary support from the Quebec Heath Research Funds (Junior 2 and Senior Scholar respectively)

### Competing Interest Statement

The authors have declared no competing interest.

### Funding Statement

No external funding was received for this study. Krista Best and Francois Routhier received salary support from the Quebec Heath Research Funds (Junior 2 and Senior Scholar respectively).

### Author Declarations

This study was exempted from ethical approval by the Research Ethics Board in rehabilitation and social integration of the Centre Integre Universitaire de Sante et de Services Sociaux de la Capitale-Nationale (#2025-32390).

